# INTRANASAL APPLICATION OF *LACTOCOCCUS LACTIS W 136* BACTERIA EARLY IN SARS-Cov-2 INFECTION MAY HAVE A BENEFICIAL IMMUNOMODULATORY EFFECT: A PROOF-OF-CONCEPT STUDY

**DOI:** 10.1101/2021.04.18.21255699

**Authors:** Leandra Mfuna Endam, Cécile Tremblay, Ali Filali, Martin Yvon Desrosiers

## Abstract

**Justification:** Stimulation of early innate anti-viral responses during the early phase of SARS-COV-2 infection oxygen may improve evolution of illness and late pulmonary complications. This may be possible using a TLR agonist such as a probiotic bacterium possessing desirable immunomodulatory properties.

**Method:** We performed a non-contact, open-label, prospective randomized clinical trial comparing intranasally applied *Lactococcus lactis W136* with saline irrigation alone in patients within 96 hours of diagnosis of SARS-COV-2 infection not requiring supplemental oxygen.

**Results:** Twenty-three of a planned forty participants aged 18-59 without comorbidities were recruited. Irrigation with intranasal *L lactis W136* twice-daily for fourteen days of was associated with a nasal response characterised by increase in the symptom of Facial and Throat pain/discomfort, and with a lesser severity in symptoms of i) Fatigue ii) Olfactory dysfunction and iii) Breathlessness. Safety and tolerability were good, with no acute infections or severe deteriorations.

**Interpretation:** Facial and throat pain may correspond to postulated mechanism of action corresponding to activation of innate defences with antiviral effects and may explain the potentially protective effects seen. Intranasal *L lactis W136* irrigations may thus represent a potentially inexpensive, safe, and easily scalable non-antigen based therapeutic for the continuing global SARS-COV-2 pandemic.

**Data availability statement:** Data is available on request from the senior author, Dr Desrosiers: *martin-yvon*.*desrosiers*.*med@ssss*.*gouv*.*qc*.*ca*

**Funding:** This work was supported with internal funds from the Desrosiers laboratory at the Centre de Recherche du Centre Hospitaller de l’Université de Montreal (CRCHUM).

**Competing Interests:** Dr Desrosiers holds equity in Probionase Therapies inc., Which commercialises *Lactococcus lactis W136* for chronic rhinosinusitis.

## MANUSCRIPT

### INTRODUCTION

SARS-COV-2 is an infectious disease which is both extremely contagious and morbid (1). On a more worrisome note, emerging novel variants may be even more contagious, and may hamper the effectiveness of vaccine-based and antibody-based therapies (2). This suggests that the current SARS-COV-2 pandemic may continue to play havoc with the physical and mental health of our global society for some time to come, underlining the urgent need for new approaches. Current approaches focus on prevention of infection via social distancing, use of personal protective equipment, and vaccination, and management of late hyper-inflammatory complications. Treatment of established infections not requiring oxygen remains supportive, with only antiviral antibodies available to reduce SARS-COV-2 viral burden. However, this approach is increasingly questioned with the rise of the variants that may render single-antibody solutions obsolete and ‘cocktails’ of multiple antibodies too expensive and difficult to manufacture.

Modulation of early inflammatory responses crucial to the later phases of infection to produce more appropriate initial and late responses to the virus may represent a novel approach. While mortality in hospitalised pateints appears to be related to a hyper-inflammatory state complicating existing comorbidities (3, 4), it is increasingly suspected that the late inflammation observed in with SARS-COV-2 is a function of impaired immune signalling and effector responses during the early phase of infection (5). Innate immune responses trigger simultaneous activation of the NFkB complex, leading to production and secretion of several chemokines, and the activation of anti-viral genes collectively termed the ‘Interferon Signalling Gene complex’ (ISG) responsible for antiviral activity. (6) Collectively, activation of the ISG culminates in generation of multiple interferons, with certain interferons expressed only by epithelial cells. SARS-COV-2 interferes both with the early events of innate signalling and also with interferon responses (7, 8). This is associated with a delay in innate immune responses and reduced or delayed interferon production (9). It has been suggested that this renders the host response insufficient to contain virus replication within the upper respiratory tract and contributes to its subsequent spread to the lung (10).

In the absence of early innate signalling which coordinates cellular proliferation and activation, subsequent immune responses can be inappropriate and contribute to the development of a hyper-inflammatory state, which in its most severe form can lead to the highly lethal ‘cytokine storm’, or to prolonged post-SARS-COV-2 symptoms (11). This hyperinflammatory state is associated with a depletion in bacterial diversity and quantity in the oropharynx and trachea, which is believed to contribute to the severity of the underlying process (12). Additionally, it is believed that this leads to ineffective adaptive immune responses at the mucosal level. This has been demonstrated by comparative studies of serum and mucosal fluid antibody levels post-infection, where robust serum antibody responses can be seen after infection, but that these are only infrequently accompanied by a similar response at the mucosal level. (13) This is believed to be a function of insufficient stimulation for antibody production at the level of nasal mucosa, which may leave previously infected individuals vulnerable to re-infection. These observations may help explain how the absence or low levels of symptoms during the early phases of SARS-COV-2 19 infection may be associated with a more severe evolution, the development of complications, and even possibly defective protective mucosal antibody responses. Thus, the prolonged asymptomatic silent period following initial SARS-COV-2 infection not only contributes to its ease of spread between individuals but may also dictate the degree of disease severity.

Given the dysfunctions described in immune signalling and interferon production, attempts to reproduce or stimulate natural anti-viral defences, using non-antibody-based strategies, are under assessment. Intranasal application if interferon alpha has previously been explored as prophylaxis and treatment for rhinovirus and influenza infection (14). Its effect is believed to be via induction of the ISG complex with activation of antiviral defences, but toxicity has limited its clinical use. Interferon-lambda is less toxic and has been proposed as prophylaxis or treatment (5), via a similar mechanism as postulated for alpha interferon. As an alternative to interferon-alpha or lambda, it may also be possible to generate antiviral responses by stimulating detuned innate immunity, using a strong TLR stimulus applied intranasally early in the course of disease. This has been successfully demonstrated in the hamster model of SARS-COV-2 infection using a TLR3 agonist (16). Intranasal application of the TLR3 agonist Poly I:C after SARS-COV-2 infection was associated with a reduced level of virus in lung tissue, in a similar fashion to that seen with intranasal interferon-α A/D. This supports the hypothesis that antiviral activity can be obtained by supra-physiologic stimulation of the innate system.

We believe we can stimulate innate immune activation by administering intranasally a probiotic bacterium already commercialised for intranasal application. High levels of bacterial TLR motifs on the bacterial surface and in cytoplasm could potentially activate innate immune signalling and activate a cascade culminating in antiviral activity. This should lead to more rapid pathogen clearance and condition the adaptive immune system to develop a coordinated mononuclear response and antibody generation in the mucosal compartment, with potentially protective effects on the lung. This desirable characteristic needs to be balanced with a favorable side effects profile, as activation should not generate excessive pro-inflammatory responses locally or systemically. *The Lactococcus lactis W 136* bacteria already in use for CRS management possesses particular properties rendering it desirable for this application, both as a TLR agonist implicating TLR’s 2, 4, 6 on its surface and an abundance of TLR3 moieties in its cytoplasm. (17) Additionally, it has properties to increase microbiome diversity (18) and the presence of the beneficial bacterial species such as *Dolosigranulum Pigrum* (19) which might prove beneficial to patients at later stages of the disease. We explored the hypothesis that administration of probiotic *Lactococcus lactis W136* bacteria directly to the upper airway via irrigation to patients receiving a diagnosis of SARS-COV-2 will reduce symptom severity and disease in patients with SARS-COV-2 disease not requiring ICU admission or intubation. This strategy could have role in treatment, and possibly in prevention of infection in high-risk populations.

To this end, we performed a “proof of concept” clinical trial to compare the safety and validity of intranasal irrigation with L. lactis W136 bacteria applied intranasally n in recently diagnosed SARS-COV-2 infection not requiring supplemental oxygen. In order to maximise safety, healthy younger participants free of comorbidities were recruited, and non-contact techniques to protect research personnel were used for trial performance.

## METHOD

### Study Design

We performed a non-contact, open-label, prospective randomised clinical trial comparing intranasally applied *Lactococcus lactis W136* with saline irrigation alone in patients within 96 hours of diagnosis of SARS-COV-2 infection. The trial took place from July 1, 2020 to February 15, 2021 at the Centre Hospitalier de l’Université de Montréal (CHUM). Approval was obtained from Health Canada (Health Canada registration number: 249512) and the CHUM Institutional Review Board and Ethics committee (Registration No. 20.012) prior to trial performance. Informed consent was obtained from all participants prior to inclusion in the study. This study was listed on clinicaltrials.gov (ClinicalTrials.gov Identifier: NCT04458519).

Subjects were recruited via advertising in SARS-COV-2 clinics, the hospital Intranet and closed-circuit hospital TV network, and radio, newspaper and social media. This was complemented by coverage of the trial locally on network television and newspapers. No financial incentive was provided to subjects for participation. All measures were obtained and processed ensuring patient data protection and confidentiality. Complete patient recruitment and enrolment flow chart is presented in Figure 1 as per CONSORT standards. (20)

**Figure 1.**
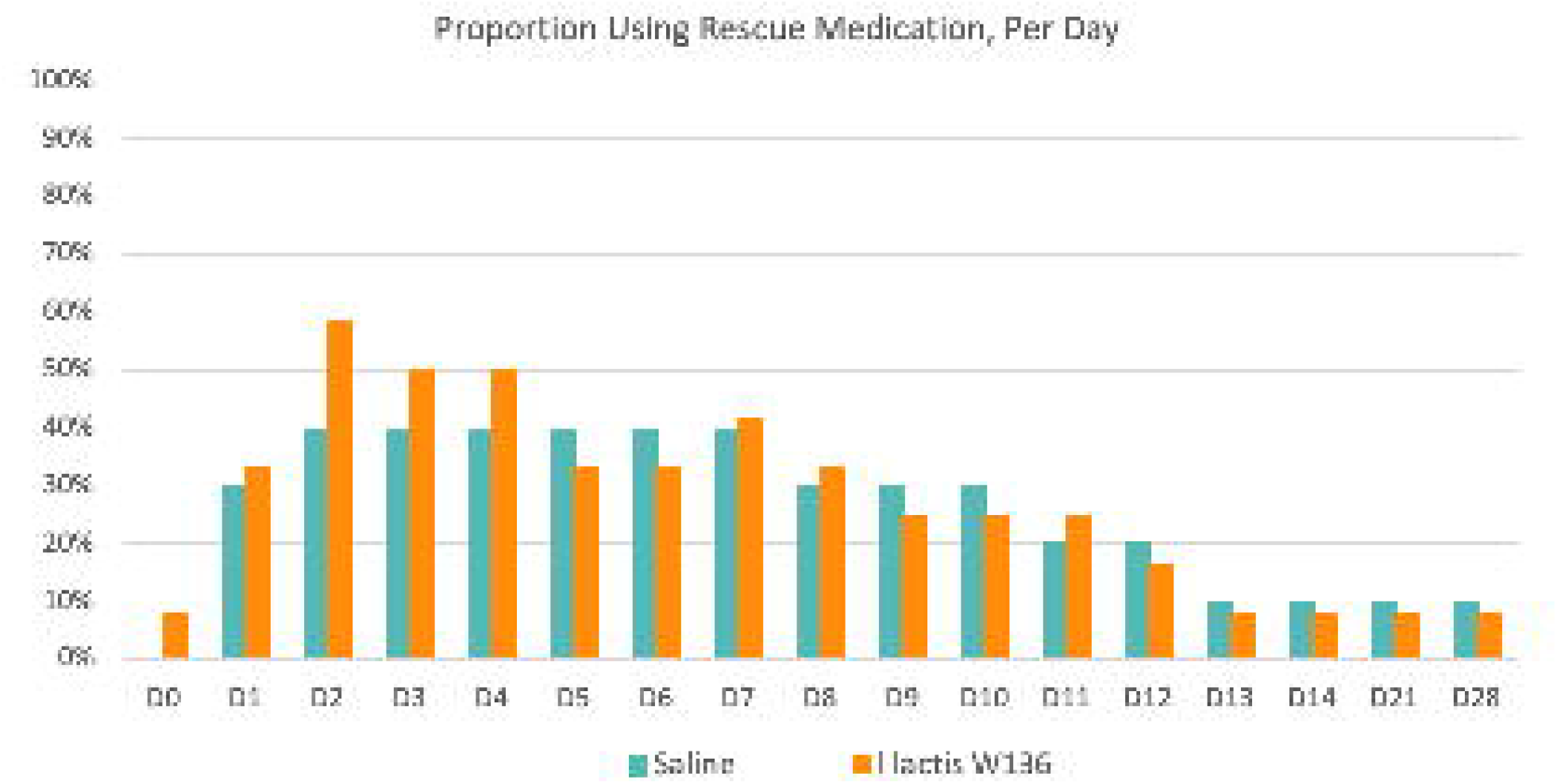
CONSORT diagram showing details of trial.

Patients 18-59 years having received a recent positive SARS-COV-2 PCR test result (within the past 96 hours) were eligible for participation if they met the other inclusion and exclusion criteria. All tests were PCR based on nasal swabs collected and processed in Government of Quebec-run facilities performed according to established Quebec Public Health protocols. All PCR tests were processed in government facilities. Infected patients usually received their diagnosis with 24-36 of testing, with results communicated to them directly via telephone. The trial protocol is summarised in **Figure 1**. Healthy males and females aged 18-59 years without comorbidities were recruited. Subjects were required to present proof of a positive PCR diagnosis of SARS-COV-2 infection made within less than 96 hours by the central provincial diagnostic laboratory. Patients had to have a temperature at entry of less than 38.0 °C and no requirement for supplementary oxygen, nor could they be hospitalised. Patients had to be able and willing to perform nasal irrigation, had to provide consent, and needed to be able to communicate with the study team by phone, text or email. <u>Exclusion criteria:</u> Patients with pre-existing conditions or demographic features placing them at increased risk of complications from SARS-COV-2 infection were not included in this study. This included hypertension, cardiovascular disease, and diabetes, cystic fibrosis, asthma, COPD, bronchiectasis, and individuals with immunosuppression from primary immune deficiencies, chemotherapy or immune suppressing medications. Pregnant or breastfeeding women, or women unwilling to practice contraception were excluded.

### Intervention

Treatment consisted of twice daily nasal irrigations with either a buffered isotonic solution containing 2.4 Billion CFU of *Lactococcus lactis W136*, or buffered isotonic saline alone, for a period of 14 days. Pre-measured individual sachets containing treatment were reconstituted in 240 ml of clean water. and nasal irrigation then performed by the patients using a NeilMed SinusRinse irrigation bottle. This was followed by a 2-week observation period where patients received no treatment but continued to record their symptoms.

Subjects received delivery of study material at their place of confinement and were advised to perform nasal irrigation alone in a closed room to minimise potential aerosol generation and dispersion to those in their household. Interviewing for details of medical history and SARS-COV-2 symptom development was done by telephone or email, with follow-up performed remotely by phone, text message or email as desired by the patient. Intranasal or allergy medications were allowed to be continued if they were used prior to the trial, including oral antihistamines or intranasal corticosteroid for seasonal allergic rhinitis or homeopathy for any purpose. Use of systemic or topical antibiotics, topical nasal decongestants, or oral corticosteroids was forbidden. Rescue medications or treatments were permitted for symptomatic relief of non-specific symptoms of SARS-COV-2 19 infection. These included acetaminophen, non-steroidal anti-inflammatories, oral decongestants, use of a humidifier, and aromatherapy. All medication and treatment use were recorded in the patient diary.

### Monitored parameters

Assessments were performed remotely and thus relied upon non-invasive parameters which could be recorded by the patient. No nasal examination or brushings were performed. Given the lack of validated instruments to measure SARS-COV-2 symptoms, we opted to assess all of the symptoms accepted as reported in SARS-COV-2 infection at the time of trial design. These included systemic, nasal, pulmonary and digestive symptoms. Intensity of symptoms was assessed using a visual analogue scale (VAS) as we believed it offered a better dynamic range of response than a three-point ordinal scale. Intensity of individual symptoms was thus graded using a 100mm visual analogue scale to yield a VAS score that could range from 0 (Not troublesome) to 100 (Worst thinkable troublesome). The VAS score was interpreted as follows: Mild = VAS 0 to 30, Moderate = VAS >30 to 70 and Severe = VAS >70 to 100. Symptoms were assessed daily for fourteen days, and once weekly for the following 14 days. Patients recorded their scores using paper diaries, reminded by prompts from study staff. Patients monitored their temperature daily with digital thermometers supplied with study materials. Symptoms warranting urgent consultation were taken from the Government of Quebec website and reproduced in the consent form for patients use. Patients were urged to consult rapidly at emergency if severity criteria developed.

### SARS-COV-2 mitigation strategies

Recruiting and trial performance strategies to minimise risk of exposure to SARS-COV-2 virus and patients’ moments outside their confinement area were employed. These reflect the high level of concern over the risk of SARS-COV-2 transmission and infection at that time. At the time of study design, risks and mortality from SARS-COV-2 infection were presumed quite high, and the mode of transmission had not yet well been elucidated, making contact presumed quite risky. Strategies were thus deployed to minimise to reduce in-person contact between infected patients and study staff, and also to avoid patient movement outside the home to avoid increasing contagion. This led to study performance using remote monitoring, with in person visits reserved only for the delivery of study material. Patients were recruited and consented was obtained verbally over the telephone. Assessments relied upon non-invasive parameters recorded by the patient on a paper diary furnished at the initiation of the study. All assessments were performed remotely and were prompted by daily reminders by phone or e-mail.

### Strategies for reducing bias

This trial was single blinded and open label. In order to minimise bias, treatment was allocated at the time of delivery, when an envelope containing the code was opened by the staff performing deliveries. Delivery staff had no further contact with patients.

Study staff in initial and follow up contact with pateints were kept blinded as to assignment. Any question or medical issue that could identify treatment was to be directed to one of the Investigators. Patients were able to identify their treatments, so it was emphasised that both treatments were active treatments for nasal and sinus symptoms and should thus both offer a theoretical benefit to patients. In order to ensure that dissimilar packaging and box labelling did not influence patient impressions, a similar presentation was ensured for both products by removing materials from their original packaging and repacking them in a plain box inscribed only with the study name and a number code PROBCO # 000XX) which contained the rest of study materials.

## STATISTICAL ANALYSIS

Randomisation was performed using a random number generator in blocks of twenty to ensure equal distribution of treatments. Efficacy analyses were conducted according to the treatment assigned. Results were analysed for intensity of individual symptoms and the proportion of patients with moderate or severe symptoms (over 30) for each individual symptom at each timepoint. Symptom score comparisons between the two arms at each time point were performed using the non-parametric Wilcoxon test. Proportions of subjects with daily symptom intensity moderate/severe (VAS score > 30) for each symptom were compared using Fisher exact test. All tests were performed two-sided, using a significance threshold of p < 0.05. Statistical analyses were performed using the R (version 4.0.3) statistical open-source software (*https://www.R-project.org*)

**Estimate of power** was necessarily approximate, given the multiple unknowns surrounding this evolving infectious disease. We determined that with twenty patients in each arm, we would be powered to detect a major effect size if one was present. We expected at least 90% of the study population to be able to successfully perform irrigation, as clinical experience (unpublished data) suggested that less than 5% of individuals would be unable to perform it.

We also expected a less than 5% drop out due to excess severity of SARS-COV-2 19 infection, as comorbidities and age groups associated with poor outcome were excluded. We thus estimated that 18 subjects in each group would complete their treatment and be suitable for analysis. With this number we will be able to detect large size effect with a power of 80% and a type I error rate of 0.05. Assuming a standard deviation of 50% of the numerical result, this would yield a 95% power to detect a 50% difference in outcomes, and 82% power to detect differences of 40%. Thus, in this small exploratory trial, we expected to be able to capture major differences in response if they were present, generating information which can serve to develop larger, better powered trials.

### Imputation strategy for missing values

Given the unpredictable nature of SARS-COV-2 infection at time of study design, no *a priori* assumptions were made regarding disease evolution and there was no imputation strategy used for missing values. This forced us to perform a per-protocol assessment, as opposed to ‘Intent to treat’ strategy, as the single withdrawn subject did not maintain their symptom diary and did not remain in contact with study staff, so no values were available apart from initial scores solicited during the recruitment telephone evaluation.

## RESULTS

Twenty-three patients were randomised to receive study treatment. One patient (saline-only) was withdrawn from the study at time of delivery of materials because of investigator concerns regarding apparent severity of patients’ dyspnea and cough and was instead directed to the emergency room for urgent assessment. The remaining 22 patients completed the trial.

Demographics at study entry reflected the study population (Table 1). Patients were younger, and as per protocol did not have comorbidities. There was a greater participation of females, but this was evenly distributed between groups. Symptoms at time of study entry (Table 2) varied between patients, but no consistent pattern could be discerned. Patients presented an average of 8.7 symptoms, with an average 4.6 symptoms reported at moderate / severe intensity. Most severe symptoms at this initial timepoint were Fatigue and Limitation of activities, with average scores reported at 48.4/100 and 47.5/100, respectively. These were also the most frequently reported symptoms, with Fatigue present to some degree in 95% of subjects, and Limitation of activities in 86%. Next most severe symptom was impaired sense of smell and/or taste, with an intensity of 41.4/100. Despite its frequent reported association with SARS-COV-2, impairment of smell was reported by only 64% of subjects. This nevertheless corresponded well with responses given in telephone interviews during the recruitment process, where 60% of subjects complained of this symptom after prompting. Other symptoms were present in various combinations and variable intensities. The least frequently reported symptom was vomiting, which was reported by only one subject at baseline. There were no statistically significant differences in severity of symptoms observed at baseline, and the proportion of pateints in each group with moderate / severe symptoms was similar.

**Table 1.** Study demographics. More women than men participated but were equally distributed between the two treatment groups. As per protocol, pateints were between 18-59 years of age and free of comorbidities.

|  |  |  |
| --- | --- | --- |
| **Age in years (mean ± SD)** | | 30.4 (±9.1) |
| **Gender (n) (%)** | Male | 30% |
|  | Female | 70% |
| **Ethnicity (n) (%)** | White | 17 (75%) |
|  | Hispanic | 3 (17%) |
|  | Arabic | 1 (8%) |
| **Smoking history (n) (%)** | Current | 1 (8%) |
|  | Former | 6 (26%) |

**Table 2.** Symptoms at time of study entry. Symptoms are presented according to the frequency the symptom was reported by subjects. While fatigue is reported by almost all participants, an impaired sense of smell is reported by only half the subjects.

| **SARS-Cov-2 Symptoms at Study Entry** | **Proportion reporting** | **Average Intensity** |
| --- | --- | --- |
| Fatigue | 95% | 48.4 |
| Reduction of usual activities | 86% | 47.5 |
| Loss of sense of smell and/or taste | 55% | 41.4 |
| Headache | 82% | 35.0 |
| Musculoskeletal pain | 82% | 34.3 |
| Loss of appetite and nausea | 64% | 28.4 |
| Pain/ pressure in face | 64% | 27.7 |
| Cough with or without mucus (phlegm) | 77% | 21.8 |
| Shortness of breath with or without wheezing | 68% | 21.1 |
| Chills | 41% | 16.4 |
| Sore throat / Difficulty in swallowing | 32% | 14.1 |
| Diarrhea | 41% | 13.0 |
| Vomiting | 0% | 0.9 |

Symptoms of SARS-Cov-2 infection evolved over the course of the study. Several symptoms trended to increases over the Day 3 to Day 7, then showed gradual reductions over time (Supplemental Figure S1). This was accompanied in most instances by an increase in the proportion of subjects reporting moderate or severe intensity of symptoms (Supplemental Figure S2). The symptom presenting the greatest maximal average intensity during the trial was Impairment of Sense of Smell or Taste and was accompanied by an apparent increase in the proportion of patients reporting moderate and severe intensity of this symptom.

Treatment was well-tolerated and could be performed by all patients. The nasal irrigation technique was successfully mastered by all patients, and compliance with treatment was excellent, with planned irrigations performed 96.8% of the time (96.4% saline-only, 97.1% *L lactis W136*; p=NS).

Treatment with *L lactis W136* was associated with differences in certain symptoms. Intranasal irrigation with *L lactis W136* was associated with a lesser proportion of patients presenting moderate/severe symptoms of fatigue, impairment of sense of smell, and sensation of breathlessness, and by an increased proportion of subjects with moderate/severe Facial pain or Sore throat. The time course of each individual symptom is presented in **Figure 3** and the proportion of subjects with moderate or severe symptoms at each time point is presented in **Figure 4**.

**Figure 2.**
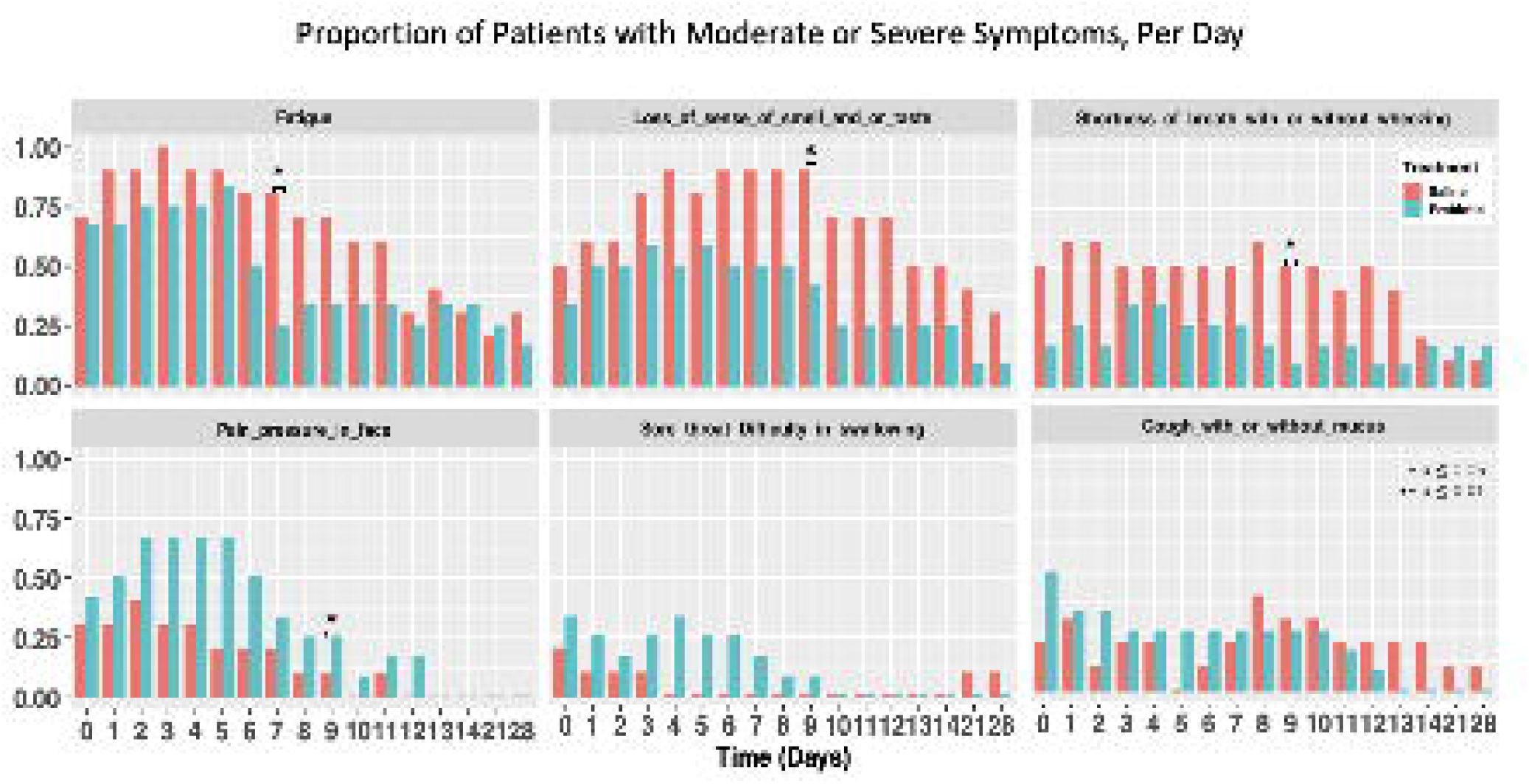
Clinical trial protocol. Following recruitment, subjects with recent SARS-CoV-2 diagnosis were randomised to 14 days of BID intranasal irrigation with *L lactis W136* 2.4 CFU solution or saline only, followed a by a further 14-day observation period. No physical contact between subject and study staff was required during the study.

**Figure 3.**
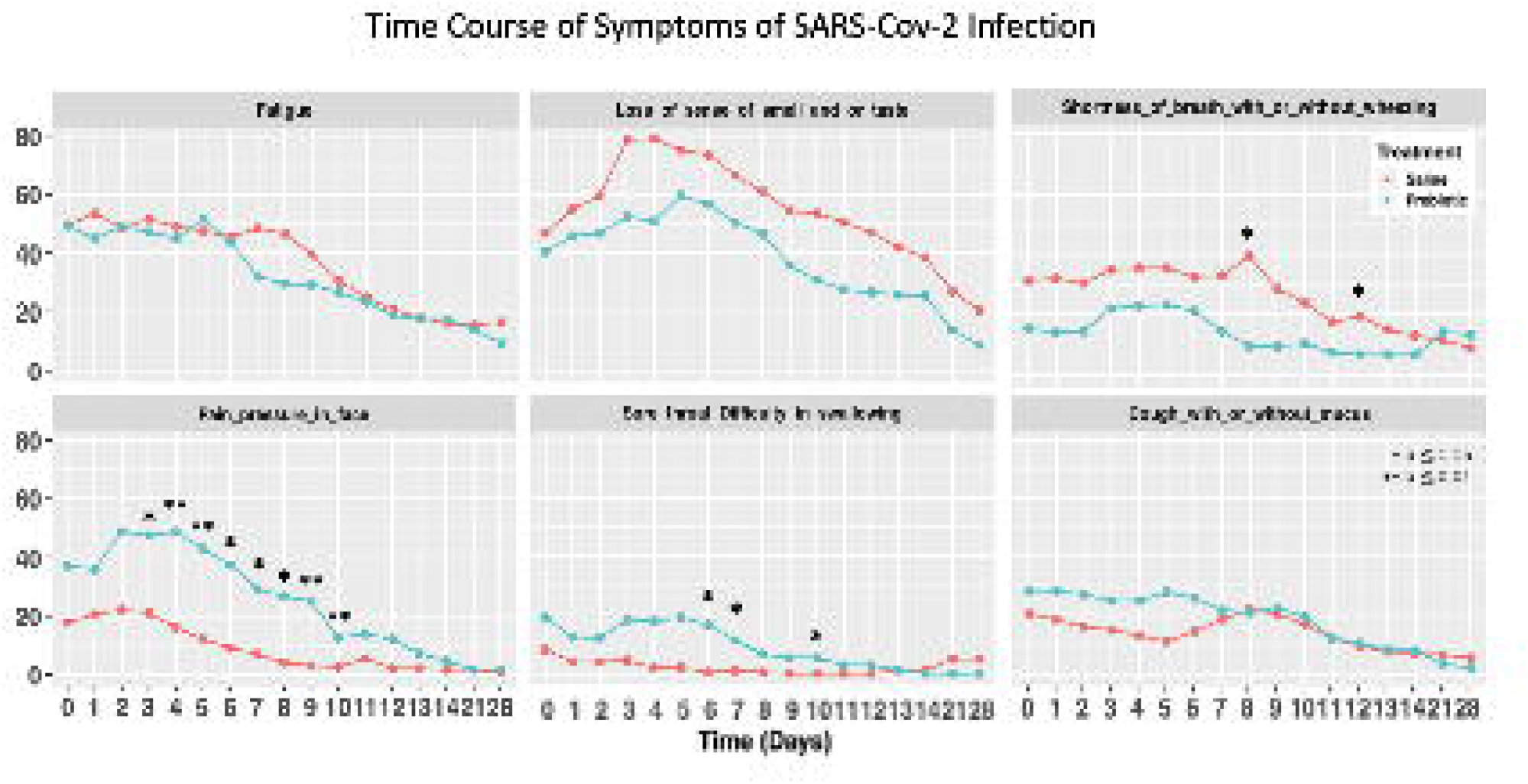
Symptom intensity over the course of the study for symptoms impacted by treatment. A signal is seen for increased Facial Pain and Discomfort in the *L lactis W136* treated group, however symptoms of Fatigue, Disturbance of Smell, Shortness of Breath trends towards a less severe course of illness in the *L lactis W136* – treated group. Interestingly, an apparent increase in cough is seen later in disease course in the saline-only group and appears to coincide with the later increase in SOB in the saline group. (*< p 0.05; **< p 0.01)

**Figure 4.**
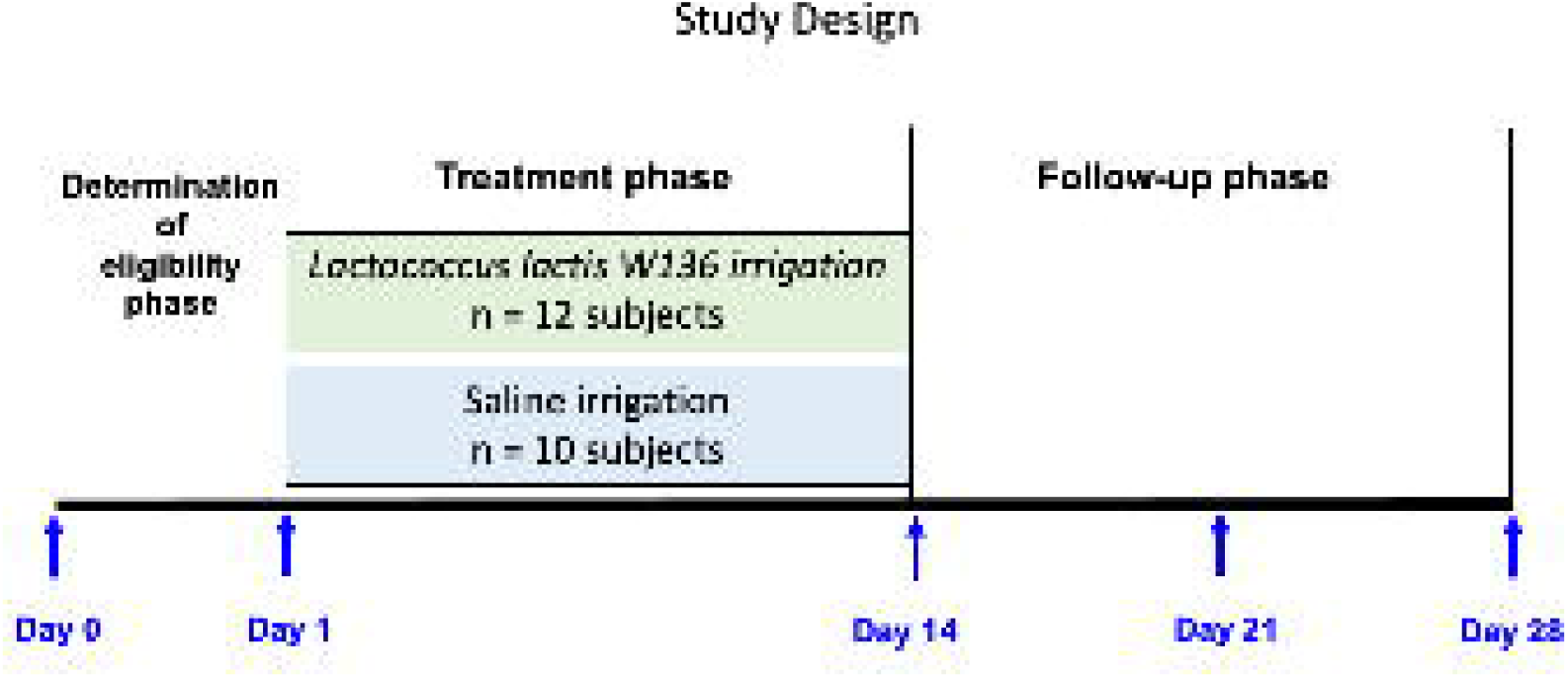
Proportion of subjects with daily symptom intensity moderate/severe (VAS score > 30) for symptoms impacted by treatment. The proportion of subjects with Facial pain is slightly greater, but on one day only. Otherwise, despite similar baseline incidences, a smaller proportion of subjects treated with *L lactis W136* report moderate/severe symptoms of Fatigue, Impairment of sense of smell, and Shortness of Breath during the course of infection than do those in the saline group. (*< p 0.05; **< p 0.01)

The proportion of subjects with fatigue of moderate/severe intensity was significantly lesser on Day 7 in the *L lactis* W136-treated group (saline-only: 50.0%, *L lactis* W136: 32.1%; p=0.02). There was also a tendency to lesser intensity of Loss of Sense of Smell in the group treated with *L lactis* W136, with reduced proportion of patients with moderate/severe Loss of Sense of Smell on Day 9 (p=0.03) and trends towards lesser severity throughout Days 4-11.

The proportion of patients with moderate/severe Shortness of breath (SOB) at baseline was not significantly different in saline-only compared to *L lactis W136* groups, however SOB appeared to evolve differently in saline and *L lactis W136* - treated groups. At days 8 and 12, SOB was significantly (p=0.02 and p=0.04 respectively) lesser in the *L lactis W136*-treated group. Moderate/severe symptoms of SOB were also reported by a lower proportion of subjects in the *L lactis W136* group on Day 9, with a borderline significance (p=0.05). Interestingly, the symptom of Cough appeared to follow a similar pattern as SOB, with a trend to a similar late increase in intensity and proportion of pateints with moderate or severe symptoms of Cough in the saline-only group.

Treatment with *L lactis W136* was associated with significantly increased in the symptom of Facial Pain/Pressure through Days 3-10 (all p≤ 0.05). While the two groups showed a similar proportion of patients with moderate/severe symptoms at baseline, moderate or severe symptoms were reported by a higher proportion of subjects in the *L lactis W136* group on Day 9 (p=0.05). The symptom of sore throat followed a time course similar to that of facial pain, but of more modest intensity.

Slightly more than half the subjects still exhibited symptoms of any intensity at study end (Saline: 58.3%, *L lactis W136*: 66.7%). At least one of these symptoms was of moderate/severe intensity in 40% of saline-treated subjects vs. 33.3% of the *L lactis W136* study group. In telephone follow up at one month post study,, persistent symptoms were reported by 30% (saline) and 25% (*L lactis W136*) of subjects, however there were no persistent symptoms of Fatigue, Anosmia, or Shortness of Breath in any of the subjects, and no differences between study groups.

### Rescue medication

**(Figure 5)** Rescue medication was used at some point by 75% of the subjects. This consisted mainly of acetaminophen and ibuprofen for pain relief. The pattern of use of rescue medication closely followed the symptoms of body, facial and throat pain. No difference in rescue medication use was seen between groups.

**Figure 5.**
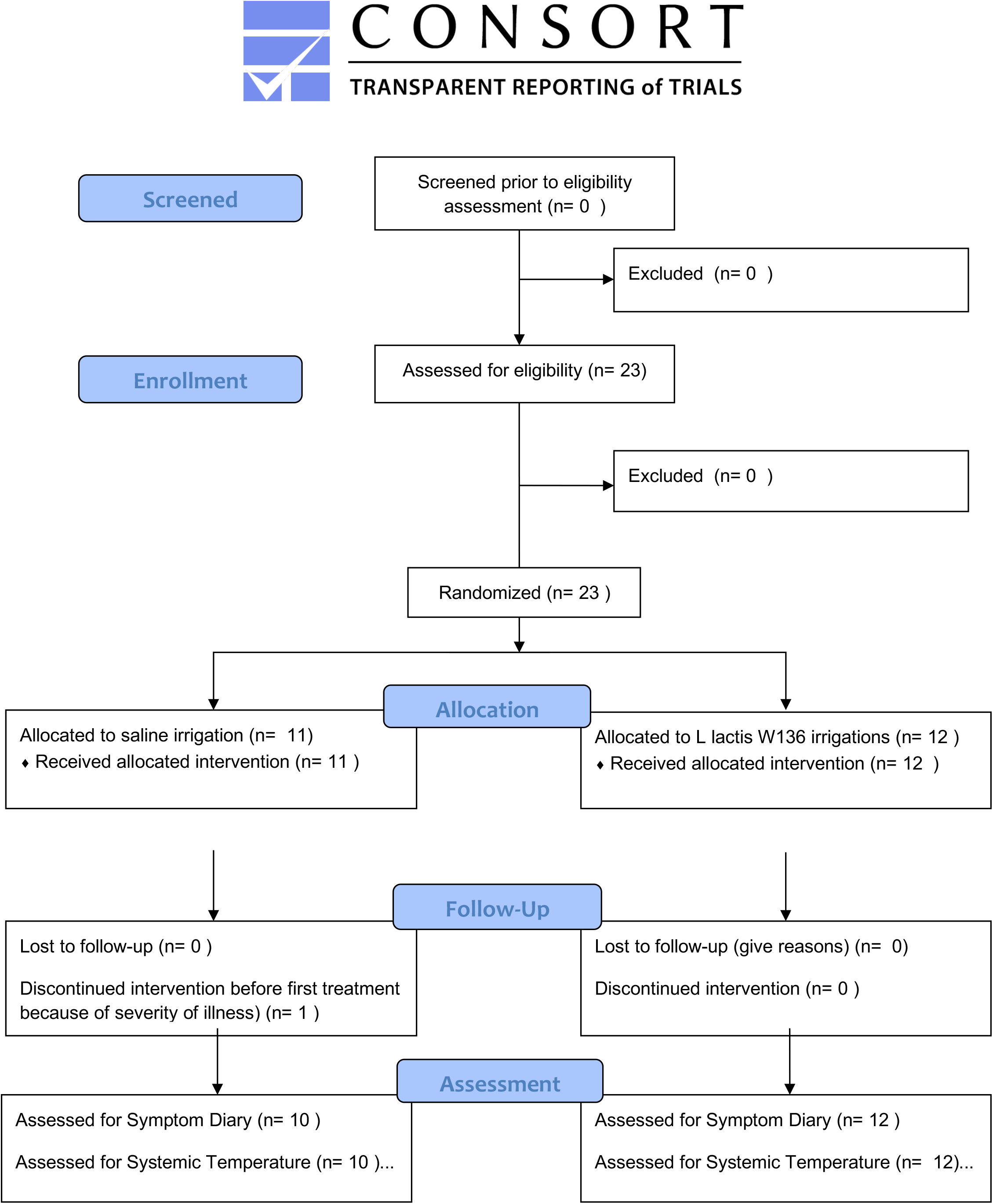
Rescue medication use during the course of the study. Similar rates of use are observed, save for a trend toward increased use in the *L lactis W136* treated group during the early days where facial and throat pain were maximal.

## ADVERSE EVENTS

Treatment-specific adverse events were rare and evenly distributed between groups (Table 3). None of these was sufficiently severe to suspend treatment(s), interrupt study participation or cause the patient to seek medical attention. No serious adverse events were noted. No episodes of acute sinusitis or acute otitis media developed during treatment. There were no episodes of nasal bleeding reported during the study period in either group.

**Table 3.** Adverse events. All events reported by subjects in their study diaries that are not disease related are reported in terms of percentages.

| **Number reported (n) (%)** | **Saline** | ***L lactis W 136*** |
| --- | --- | --- |
| Migraine | 1 (10%) | 1 (8,3%) |
| Fever | 0 | 3 (36%) |
| Insomnia | 1 (10%) | 1 (8,3%) |
| Nausea | 0 | 1 (8,3%) |
| Cold sore | 0 | 1 (8,3%) |
| Ear pain | 0 | 1 (8,3%) |
| Tingling in back of nose | 0 | 1 (8,3%) |
| Dizziness | 0 | 1 (8,3%) |
| Metallic taste in mouth | 0 | 1 (8,3%) |
| Body ache | 1 (10%) | 0 |

## DISCUSSION

Intranasal probiotic therapy is a novel immune modulating therapy which has not yet been explored in SARS-COV-2 infection (21) We present the results of a “proof of concept” clinical trial assessing the impact of intranasal administration of live *L lactis W136* administered within 96 hours of diagnosis on the course of PCR-documented SARS-COV-2 infection over a 28-day study period. This small scale, open label trial suggests that intranasal administration of *L Lactis W136* produces an effect on the upper respiratory passages and pharynx, and may be accompanied with lower symptoms of Fatigue, Loss of sense of smell, and Shortness of Breath.

The strongest signal observed was for Facial discomfort, which was significantly greater in the *L lactis 136* group. While this might be considered to represent an adverse event, we suspect instead that this represents a treatment-related effect, as this is consequent with the postulated mechanism of action whereby *L lactis 136* induces an inflammatory response in the upper respiratory passages and pharyngeal areas as part of its effect. The magnitude of this response was not anticipated. In previous trials in chronic sinusitis (18) and in heathy normosmic subjects (*data on file*), participants had anecdotally reported a ‘tingling’ sensation reported as facial discomfort or pain on first administration but had never reported facial pain as more than “mild” and never required rescue medication for relief. Intranasal symptoms may thus indicate activation of viral defences and reflect the struggle for viral clearance.

A protective effect of *L lactis* W136 was suggested by lower intensities of Fatigue, Loss of sense of smell, and Shortness of breath, suggesting a milder course of illness in this group. “Fatigue” is a non-specific symptom, but which given its frequency in the study population, might be considered to reflect the level of systemic inflammation which is characteristically present systemically despite SARS-COV-2 infection being limited to the respiratory tract. The lesser fatigue may thus reflect the events unfolding in the nose and lung. While we do not have direct objective measures of function, improvements in respiratory epithelial function are suggested by lesser Impairment of smell and Breathlessness. Olfactory dysfunction reflects injury to the sustentacular cells supporting the olfactory epithelium by the SARS-COV-2 virus (22), thus the degree of anosmia could be considered a proxy for epithelial upper airway dysfunction during symptomatic SARS-COV-2 infection. The symptom of “Shortness of breath” can be considered to reflect lower airway oxygen transferring capacity and may be considered measure of lower airway epithelial dysfunction. A lesser degree of virally induced dysfunction in both upper and lower airway were associated with intranasal *L lactis W136* treatment, suggesting improved epithelial function presumably secondary to enhanced viral clearance. While these preliminary findings obviously require further validation, they are consistent with those seen in the hamster model following both IFN-I and TLR3 stimulation. (16)

Clinical data directly supporting these concepts in humans still remains limited. Administration of interferon intranasally has been proposed since the 1980’s. (23) Early studies reported flu-like symptoms in the respiratory tract, similar to those observed by the participants receiving *L lactis W136*, but also evidence of nasal toxicity as characterised by bleeding and nasal ulcerations in a significant proportion of patients. More recently, lower-dose interferon sprays have been suggested as a means of prophylaxis against development of SARS-COV-2 infection, with regular intranasal use associated with no infections in an uncontrolled group of front-line health care workers. In a trial of exposed health care workers in China, 3000 exposed workers treated with BID intranasal interferon did not contract SARS-COV-2 infection, as compared to control populations working in similar settings (24). However, as the mechanism of action is via stimulation of inflammation, intranasal administration of IFN produces burning and discomfort in the nasal passages and throat limiting its tolerability (25). Thus, while these results are promising, interferon is an expensive product, with a narrow therapeutic window for side effects and toxicity. This may restrict its use to high-risk populations, but nevertheless underlines the potential of immunomodulation as a means of modifying disease evolution in SARS-COV-2. An alternative to administering interferon may be instead to induce its production by activating host innate signalling. The potential for success of this approach has already been suggested by the efficacy of TLR3 as an immune activator in a hamster model (16) where administration of intranasal Poly I:C, a TLR3 agonist reduced the viral load and tissue damage in the lung a fashion comparable to that seen with intranasal interferon IFNα A/D. This led the authors to suggest TLR3 agonists might represent an effective alternative to interferon for early intervention in SARS-CoV-2 respiratory disease. While there are to our knowledge no published clinical trials in humans, these approaches are nevertheless already suggested as a potential means of prevention in at risk populations such as health care workers. (26)

Safety and tolerability of nasal irrigation with and without *L lactis W136* solution was very good. Even in the context of acute illness, treatment appeared acceptable to patients and safe. Safety, an important consideration, appeared excellent. All patients receiving therapy were able to complete the entire course. There were no treatment-associated adverse effects apart from those, such as facial pain, considered to represent mechanism of action. Less than 5% of scheduled irrigations were not performed, and no patient reported difficulty mastering the irrigation technique. A concern is a single patient in *L lactis W136* group who reports maximal Shortness of Breath at days 21 and 28. However, in this instance, systemic involvement does not appear to be present, as at same timepoints where dyspnea is reported as maximal, the same individual reported a score of “0” for every other measured symptom, having completely recovered their sense of smell and manifesting no Fatigue, Limitation of activities or chills. On telephone follow up one month after end of study, this subjects’ symptoms had cleared completely. This may be considered to be reflection of the discrepancies involved between subjective questionnaires and objective assessments in other studies of SARS-COV-2 and serve to highlight the need to perform future studies in-person, with nasal examinations to collect secretions and assess toxicity, and use of home pulse oximetry to accurately assess O2 saturation.

A concern may be the effectiveness in the context of emerging variants increasing contagion and potentially reducing vaccine efficiency. variants. Our postulated mechanism of action suggests induction of non-specific anti-viral responses which should not be modified by the modification of antigen targets on the virus, as can be observed with variants, but will need to be confirmed by monitoring subject’s variant status in future studies.

### LIMITATIONS

This study has major limitations. This proof-of-concept study was designed to assess a novel therapy in the context of the early pandemic and was powered to detect only major effects, if present. While our estimates of ability to perform irrigation and study dropout rates proved accurate, recruitment was extremely challenging and we ceased the PROBCO trial early, at n=23 subjects, instead of the originally planned 40. Despite use of multiple previously effective recruitment strategies and several appearances on television and newspapers, these strategies have apparently become ineffective, and we believe that further recruitment for this trial has become unrealistic and will not be feasible in any reasonable timeframe.

The current analysis nevertheless offers important insights into possible mechanism(s) of action and suggests an effect on three important symptoms of SARS-COV-2. However, there are no objective measurements to confirm these, which is a consequence of the remote reporting format used to eliminate all physical contact between study personnel and infected subjects. Lastly, subgroups more likely to develop complications such as hypertension and diabetes were purposefully excluded from this trial. While we believe these results may be applicable to them, and the benefits possibly even more pronounced, the effects of *L lactis W136* still need to be assessed in this needy at-risk population.

Thus, while these results are promising, they require further validation, complemented by objective measures. measuring levels of virus, antibodies, and putative cytokines in blood and nasal secretions, and monitoring of physiologic parameters including oxygen saturation and smell should be incorporated into better-powered and designed future studies of *L lactis W136* which also include at-risk populations such as older subjects and those with comorbidities. If further evaluations validated this approach, *L lactis W136* irrigation could be easily and rapidly be produced and deployed on large scale, even for low-income countries. *L lactis W136* powder is non-toxic and stable at room temperature for twenty-four months, and thus easy to store, transport, and distribute. Cost of production at massive scale would be very low and production can be scaled to massive levels easily by using multiple already existing providers.

## CONCLUSION

We describe that nasal irrigation with *L lactis W136* twice-daily for fourteen days applied within 96 hours of SARS-COV-2 infection produces an increase in intensity of symptoms of facial pain and discomfort and is potentially associated with a lesser intensity of symptoms of i) Fatigue ii) Loss of sense of smell iii) Sensation of Breathlessness. Increased facial discomfort associated with administration of *L lactis W136* may represent an activation of a dormant innate immune activity and may be responsible for apparent beneficial effects.

## Data Availability

Data is available on request from the senior author, Dr Desrosiers, at

## LEGENDS FOR TABLES AND FIGURES

Supplemental Figure S1. <u>Symptom intensity over the course of the study for all symptoms assessed for all symptoms.</u>

Supplemental Figure S2. Proportion of subjects with symptom moderate/severe intensity (VAS score > 30) on each day for all symptoms.

## REFERENCES

1. Zhang H, Penninger JM, Li Y, Zhong N, Slutsky AS. Angiotensin-converting enzyme 2 (ACE2) as a SARS-CoV-2 receptor: molecular mechanisms and potential therapeutic target. Intensive Care Medicine. 2020:1–5

2. McCormick KD, Jacobs JL, Mellors JW. The emerging plasticity of SARS-CoV-2. Science 26 Mar 2021: Vol. 371, Issue 6536, pp. 1306–1308. DOI: 10.1126/science.abg4493

3. Mehta P, McAuley DF, Brown M, Sanchez E, Tattersall RS, Manson JJ. SARS-COV-2: consider cytokine storm syndromes and immunosuppression. The Lancet. 2020

4. Ong, Eugenia Ziying et al. A Dynamic Immune Response Shapes SARS-COV-2 Progression. Cell Host & Microbe, Volume 27, Issue 6, 879–882.e2

5. Lei, X., Dong, X., Ma, R. et al. Activation and evasion of type I interferon responses by SARS-CoV-2. Nat Commun 11, 3810 (2020). https://doi.org/10.1038/s41467-020-17665-9

6. Stanifer ML, Guo C, Doldan P and Boulant S (2020) Importance of Type I and III Interferons at Respiratory and Intestinal Barrier Surfaces. Front. Immunol. 11:608645. doi: 10.3389/fimmu.2020.608645

7. Ribosome-profiling reveals restricted post transcriptional expression of antiviral cytokines and transcription factors during SARS-CoV-2 infection. Alexander MR,Brice AM, van Vuren JP, Rootes CL, Tribolet L, Cowled C, Bean AG, Stewart CR. bioRxiv 2021.03.03.433675; doi: https://doi.org/10.1101/2021.03.03.433675

8. Liu Y, Yan L-M, Wan L, et al. Viral dynamics in mild and severe cases of SARS-COV-2. The Lancet Infectious Diseases. 2020.

9. Nguyen, T.H., McAuley, J.L., Kim, Y., Zheng, M.Z., Gherardin, N.A., Godfrey, D.I., Purcell, D.F., Sullivan, L.C., Westall, G.P., Reading, P.C., Kedzierska, K. and Wakim, L.M. (2021), Influenza, but not SARS-CoV-2, infection induces a rapid interferon response that wanes with age and diminished tissue-resident memory CD8+ T cells. Clin Transl Immunol, 10: e1242. https://doi.org/10.1002/cti2.1242

10. Mazewski C, Perez RE, Fish EN and Platanias LC (2020) Type I Interferon (IFN)-Regulated Activation of Canonical and Non-canonical Signaling Pathways. Front. Immunol. 11:606456. doi: 10.3389/fimmu.2020.606456

11. Habibi MS, Thwaites RS, Chang M, Jozwik A, Paras A, Kirsebom F, Varese A, Owen A, Cuthbertson L, James P, Tunstall T, Nickle D, Hansel TT, Moffatt MF, Johansson C, Chiu C, Openshaw PJM. Neutrophilic inflammation in the respiratory mucosa predisposes to RSV infection. Science, 09 Oct 2020: Vol. 370, Issue 6513, eaba9301 DOI: 10.1126/science.aba9301

12. Shen Z, Xiao Y, Kang L, Ma W, Shi L, Zhang L, et al. Genomic diversity of SARS-CoV-2 in coronavirus disease 2019 patients. Clin Infect Dis. 2020 Mar 4 [Epub ahead of print]. https://academic.oup.com/cid/advance-article/doi/10.1093/cid/ciaa203/5780800

13. Smith N, Goncalves P, Charbit B, Grzelak L, Beretta M, Planchais C, Bruel T, Rouilly V, Bondet V, Hadjadj J, Yatim N, Pere H, Merkling SH, Kernéis S, Rieux-Laucat F, Terrier B, Schwartz O, Mouquet H, Duffy D, Di Santo JP. Distinct systemic and mucosal immune responses to SARS-CoV-2. medRxiv 2021.03.01.21251633; https://doi.org/10.1101/2021.03.01.21251633

14. Kugel D, Kochs G, Obojes K, Roth J, Kobinger GP, Kobasa D, Haller O, Staeheli P, von Messling V. Intranasal Administration of Alpha Interferon Reduces Seasonal Influenza A Virus Morbidity in Ferrets. Journal of Virology Mar 2009, 83 (8) 3843-3851; DOI: 10.1128/JVI.02453-08

15. Alibek K, Tskhay A (2020) Ahead of a vaccine: A safe method of protection against SARS-COV-2 exists. Research Ideas and Outcomes 6: e61709. https://doi.org/10.3897/rio.6.e61709

16. Hoagland DA, Møller R, Uhl SA, Oishi K, Frere J, Golynker I, Horiuchi S, Panis M, Blanco-Melo D, Sachs D, Arkun K, Lim JK, tenOever BR. Leveraging the antiviral type I interferon system as a first line of defense against SARS-CoV-2 pathogenicity. Immunity, Volume 54, Issue 3, 557–570.e5 ISSN 1074-7613, https://doi.org/10.1016/j.immuni.2021.01.017.

17. Schwartz JS, Peres AG, Mfuna Endam L, Cousineau B, Madrenas J, Desrosiers M. Topical probiotics as a therapeutic alternative for chronic rhinosinusitis: A preclinical proof of concept. Am J Rhinol Allergy. 2016 Nov 1;30(6):202–205. doi: 10.2500/ajra.2016.30.4372. PubMed PMID: 28124641.

18. Endam LM, Alromaih S, Gonzalez E, Madrenas J, Cousineau B, Renteria AE and Desrosiers M (2020) Intranasal Application of Lactococcus lactis W136 Is Safe in Chronic Rhinosinusitis Patients With Previous Sinus Surgery. Front. Cell. Infect. Microbiol. 10:440. doi: 10.3389/fcimb.2020.00440

19. Brugger SD, Eslami SM, Pettigrew MM, Escapa IF, Henke MM, Kong Y, Lemon KP Dolosigranulum pigrum cooperation and competition in human nasal microbiota. bioRxiv 678698; doi: https://doi.org/10.1101/678698

20. Eldridge SM, Chan CL, Campbell MJ, Bond CM, Hopewell S, Thabane L, et al. CONSORT 2010 statement: extension to randomised pilot and feasibility trials. BMJ. 2016;355.

21. Schijns V, Lavelle EC. Prevention and treatment of SARS-COV-2 disease by controlled modulation of innate immunity. Eur J Immunol. 2020 Jul;50(7):932–938. doi: 10.1002/eji.202048693. Epub 2020 Jun 15. PMID: 32438473; PMCID: PMC7280664.

22. Cooper, K. W., Brann, D. H., Farruggia, M. C., Bhutani, S., Pellegrino, R., Tsukahara, T., Weinreb, C., Joseph, P. V., Larson, E. D., Parma, V., Albers, M. W., Barlow, L. A., Datta, S. R., & Di Pizio, A. (2020). SARS-COV-2 and the Chemical Senses: Supporting Players Take Center Stage. Neuron, 107(2), 219–233. https://doi.org/10.1016/j.neuron.2020.06.032

23. Farr BM, Gwaltney JM Jr, Adams KF, Hayden FG. Intranasal interferon-alpha 2 for prevention of natural rhinovirus colds. Antimicrob Agents Chemother. 1984;26(1):31–34. doi:10.1128/aac.26.1.31

24. Meng Z, Wang T, Chen L, Chen X, Li L, Qin X, Li H, Luo J. An experimental trial of recombinant human interferon alpha nasal drops to prevent SARS-COV-2 in medical staff in an epidemic area. medRxiv 2020.04.11.20061473; doi: https://doi.org/10.1101/2020.04.11.20061473

25. Lin F, Shen K. Type I interferon: From innate response to treatment for SARS-COV-2. Pediatr Investig. 2020;4(4):275–280. Published 2020 Dec 28. doi:10.1002/ped4.12226

26. Market M, Angka L, Martel AB, Bastin D, Olanubi O, Tennakoon G, Boucher DM, Ng J, Ardolino M and Auer RC (2020) Flattening the SARS-COV-2 Curve With Natural Killer Cell Based Immunotherapies. Front. Immunol. 11:1512. doi: 10.3389/fimmu.2020.01512

