## Supplementary figures and images for "INTRANASAL APPLICATION OF *LACTOCOCCUS LACTIS W 136* BACTERIA EARLY IN SARS-Cov-2 INFECTION MAY HAVE A BENEFICIAL IMMUNOMODULATORY EFFECT: A PROOF-OF-CONCEPT STUDY"

### Supplemental Figures S1

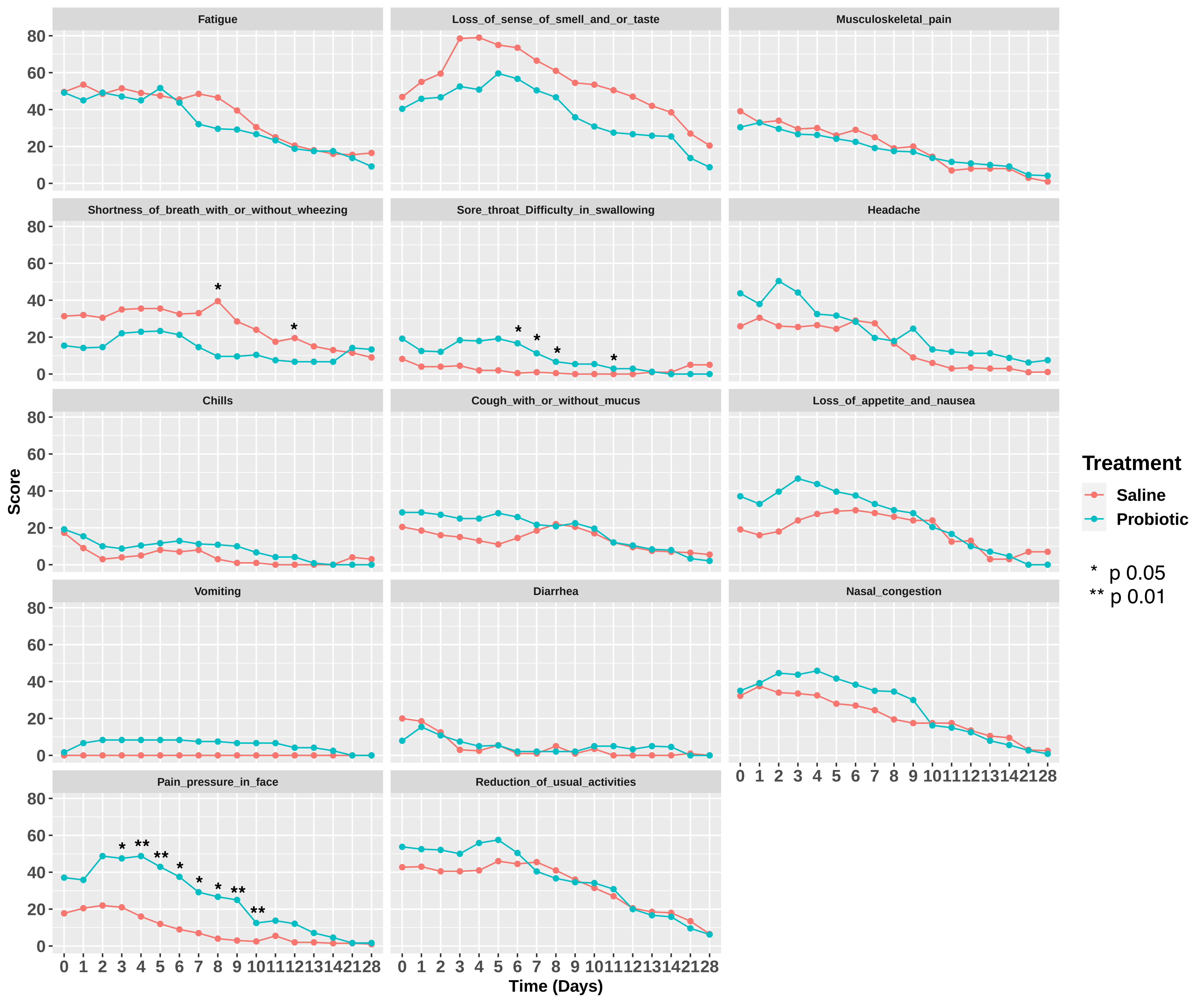

### Supplemental Figures S2

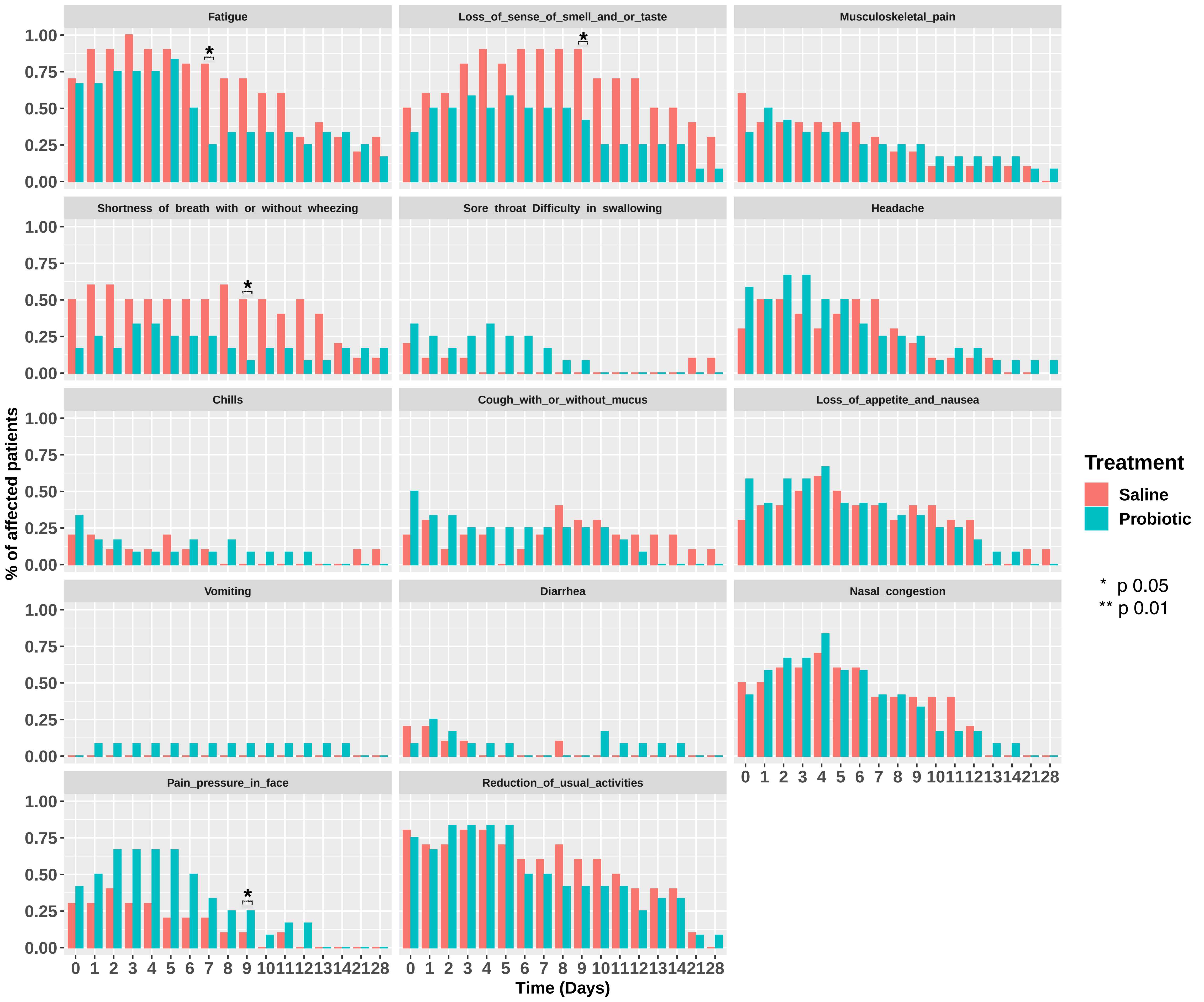
